# Volitional control of movement interacts with proprioceptive feedback in motor cortex during brain-computer interface control in humans

**DOI:** 10.1101/2024.02.26.24303289

**Authors:** Monica F. Liu, Robert A. Gaunt, Jennifer L. Collinger, John E. Downey, Aaron P. Batista, Michael L. Boninger, Douglas J. Weber

## Abstract

Vision and proprioception regulate motor output during reaching. To study the effects of sensory input on motor control, brain computer interfaces (BCIs) offer particular advantages. As part of a long-term clinical BCI trial, we implanted two 96-channel microelectrode arrays into M1 of a person who was completely paralyzed below the neck but retained intact somatosensation. Neural recordings from M1 were transformed into a 2-dimensional velocity control signal for a robotic arm using an optimal linear estimator decoder that was calibrated while the participant imagined performing movements demonstrated by a virtual arm. Once the decoder was calibrated, we asked the participant to move the robotic arm left and right past a pair of lines as many times as possible in one minute. We examined how visual and proprioceptive feedback were incorporated into BCI control during this task by providing the participant with either visual or proprioceptive feedback, both, or neither. Proprioceptive feedback was provided by moving the participant’s own arm to match the movement of the robotic arm. Task performance with vision or proprioception alone was better than when neither were provided. However, providing proprioceptive feedback impaired performance relative to visual feedback alone, unless the decoder was calibrated with neural data collected while both visual and proprioceptive feedback were provided. Providing proprioceptive feedback during decoder calibration rescued performance because it better captured M1’s neural activity during BCI control with proprioceptive feedback. In general, BCI performance was positively correlated with how well the decoder captured variance in neural activity during the task. In summary, we found that while the BCI participant was able to use proprioceptive feedback regardless of whether the decoder was trained with vision only or vision and proprioception, training the decoder with both visual and proprioceptive feedback made performance more robust to the addition or removal of visual or proprioceptive feedback. This was because training a decoder with proprioceptive feedback allows the decoder to take advantage of proprioception-driven activity in M1. Overall, we demonstrated that natural sensation can be effectively combined with BCI to improve performance in humans.

## Introduction

Sensory feedback is crucial for regulating motor output during reaching. Restoring sensory feedback to brain computer interface (BCI) users is a primary goal of neuroprosthetic research today, for both the control of action and for a restoration of somatosensation. In fact, when proprioceptive feedback is lost, people cannot make co-ordinated or accurate reaching movements in the absence of visual feedback (Ghez et al., 1995). Human studies have demonstrated that intracortical microstimulation of somatosensory cortex can provide information about tactile sensations (Fifer et al., 2022; Flesher et al., 2016; Hughes et al., 2021) and can even improve motor performance (Flesher et al., 2021). There is also interest in restoring proprioceptive input (Armenta Salas et al., 2018; Deo et al., 2021), but in order to do so effectively, we must understand how natural proprioception influences activity in motor cortex (M1) and BCI control.

Canonically, activity changes in M1 neurons precede changes in movement velocity by 100-150ms (Fetz et al., 1980; Suminski et al., 2009) and the intended velocity of movement can be decoded from the firing rates of these neurons (Georgopoulos et al., 1986; Moran & Schwartz, 1999). However, the encoding of movement velocity by the activity of M1 neurons has also been shown to vary with the posture of the arm (Scott & Kalaska, 1997; Sergio & Kalaska, 2003). Neurons in M1 also respond to sensory information, including proprioceptive (Evarts & Fromm, 1977; Suminski et al., 2010), visual (Georgopoulos et al., 1989; Pellegrino & Wise, 1993; Pellizzer et al., 1995; Zhang et al., 1997), and tactile (Schroeder et al., 2017) inputs. Responses in M1 to proprioceptive or kinesthetic inputs can lag movement by several hundred milliseconds (Evarts & Tanji, 1976; Hatsopoulos & Suminski, 2011; Suminski et al., 2009).

The postural and feedback-driven changes in M1 activity raise questions about how proprioception interacts with movement-related activity that is being decoded during BCI control. In a previous study by Suminski et al., 2010, monkeys using a BCI that was calibrated with visual feedback showed improvement in a target tracking task when proprioceptive feedback was provided via a robotic exoskeleton compared to when monkeys were relying on visual feedback alone. Furthermore, despite variation in individual neurons, proprioceptive feedback, on average, increased the firing rates of neurons in M1, indicating that proprioceptive feedback can drive M1 responses and proprioceptive information may influence BCI control.

To understand the influence of proprioceptive feedback on M1 activity and BCI performance, we examined the impact of providing proprioceptive feedback during decoder calibration and during BCI control. BCI decoders were trained with vision, or with vision and proprioception, and tested in task conditions involving a variety of combinations of visual and proprioceptive feedback while the participant controlled the movements of a robotic arm. During BCI control, the mapping between neural activity and movement was defined entirely by the decoder. This decoder was a set of weights on each channel, which can also be thought of as a vector or axis in high-dimensional neural space. In this scenario, we can examine how proprioceptive feedback influences neural activity relative to the decode axis to understand how proprioception interacts with BCI control and decoder training.

We found that proprioceptive feedback could be used for BCI control regardless of whether the BCI decoder was trained with visual feedback only, or with visual and proprioceptive feedback. However, proprioceptive feedback impaired control of the BCI trained with vision only. While proprioceptive feedback did not consistently increase or decrease the firing rates of neurons in M1, training the BCI decoder with proprioceptive feedback ensured that the decoder captured more of the proprioception-driven changes in neural activity during the task.

## Methods

These experiments were performed under an FDA Investigational Device Exemption as part of an ongoing early feasibility trial to evaluate the safety of an intracortical braincomputer interface for long term neural recording (NCT01364480). Described here is an overview of the relevant components of the experiment. Further details on surgical procedures, array implantation, and decoder training are published in Collinger et al., 2013 and Boninger et al., 2013.

### Neural recordings

Two 96-channel intracortical microelectrode arrays (4mm × 4mm, Blackrock Microsystems, Salt Lake City, UT, USA) were implanted in the hand and arm region of the left motor cortex (M1) of a woman who had been diagnosed with spinocerebellar degeneration (Collinger et al., 2013) that resulted in complete motor paralysis below the level of C4, but intact sensation. During each recording session, neural signals were recorded with a NeuroPort data acquisition system (Blackrock Microsystems, Inc., Salt Lake City, UT), and single- and multi-unit activity was identified via threshold crossings. Threshold crossings were converted to spike counts in 30 ms bins (Collinger et al., 2013). For the analyses presented here, spike counts were reprocessed into 90 ms bins and smoothed with a Gaussian kernel with a width of 180 ms.

### Decoder training

The BCI decoder was trained with neural data collected while the participant watched a virtual reality model of an arm moving automatically to 5 targets in a 2D workspace in a virtual reality environment (Virtual Integration Environment, Applied Physics Laboratory, Johns Hopkins University) (Armiger et al., 2011). While watching the visual cue, the participant was instructed to imagine moving the virtual arm to the target. Neural data and endpoint velocity of the virtual hand were recorded continuously during the calibration trials and used to train a linear encoding model that mapped the velocity in the horizontal and vertical directions to the square-root transformed firing rate of each unit. The model coefficients were fit with an ordinary least squares estimator with ridge regression (Collinger et al., 2013; Marquardt, 1970). An initial decoder for 2D (horizontal and vertical) velocity was computed using indirect optimal linear estimation (Wang et al., 2007).

In a second calibration step, the participant used this decoder to control the velocity of a robotic arm to reach to 5 possible targets in space, while the decoded output was restricted to a path reflecting the optimal reach direction (Velliste et al., 2008). A new BCI decoder was computed from the neural data and kinematics collected during this second step of closed-loop calibration. This decoder was then used for the rest of the testing session with some gain adjustment as needed based on feedback from the participant. See Collinger et al., 2013 for full details and equations.

### Line-crossing task

This study was conducted on nine days spanning a 156-day period beginning 361 days after implant. On each day, the participant used a BCI decoder to control the movements of a robotic arm (Figure 1A) (Johannes et al., 2011; Johannes et al., 2020). Two types of decoders were trained: one with visual feedback alone (V Decoder), and one in which proprioceptive feedback was provided by physically moving the participants arm to match the movement of the virtual robot arm (VP Decoder). The participant was provided with a V Decoder on five out of the nine days, and with a VP Decoder on the remaining four days (Figure 1B). Only one decoder was used each day.

**Figure 1:**
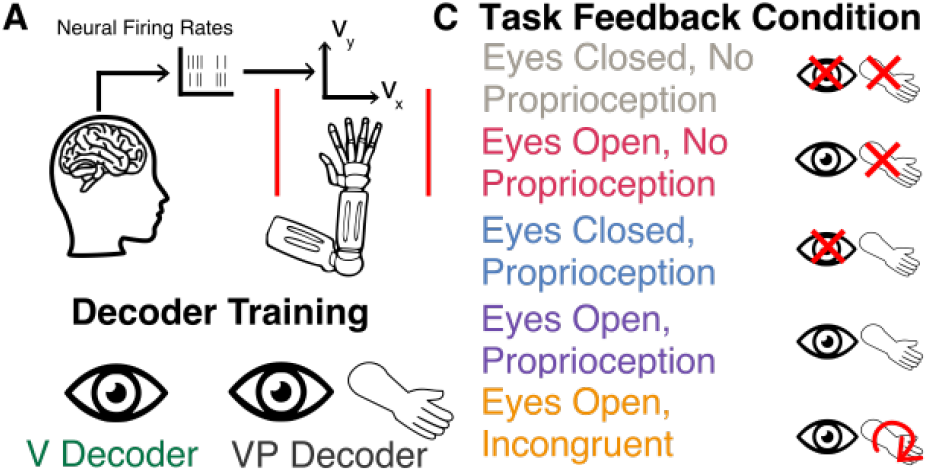
Overview of experimental setup. A: Neural firing rates, recorded from multielectrode arrays implanted in M1, were transformed into the horizontal and vertical velocity of a robot arm. The participant was instructed to move the robot arm back and forth across two lines (shown in red) as many times as possible in one minute. B: On each day, the BCI decoder was trained to map M1 neural firing rates to the velocity of the robotic arm. On 5 out of 9 days, the decoder was trained with visual feedback only; on the remaining 4 days, the decoder was trained with vision and proprioception. C: During the line-crossing task, the participant was provided with different combinations of visual and proprioceptive feedback. Each decoder training modality was paired with all task feedback conditions.

In the line-crossing task, the participant’s goal was to move the BCI-controlled robotic arm over a pair of lines near the right and left edge of the workspace. A single trial in the line-crossing task lasted for one minute, during which the participant attempted to move the robot arm leftward and rightward across the workspace as many times as possible. A pair of lines spaced 0.2m apart on the edges of the workspace served as the minimum horizontal displacement that counted as a single crossing. In each trial, the participant was provided with one of the following combinations of visual and/or proprioceptive feedback:

1. Eyes Open, No Proprioception
2. Eyes Open, Proprioception
3. Eyes Closed, No sProprioception
4. Eyes Closed, Proprioception
5. Eyes Open, Incongruent Proprioception

Four to five of each of trial types 1-4 were run on each day. On one or two trials a day, the participant was also provided with incongruent proprioceptive feedback (Eyes Open, Incongruent Proprioception), where the proprioceptive feedback was in the opposite direction relative to the movement of the robot arm (Figure 1C). In other words, as the robotic arm moved to the left, the participants own arm was moved to the right. On these trials, the participant was instructed to ignore proprioceptive feedback and focus on the visual feedback only. In total, the participant performed 25-30 trials each day. Proprioceptive feedback was provided by an experimenter manually moving the participant’s arm to match the movements of the robotic arm.

### Segmentation of kinematics and neural data

The robot arm used in this study was the Modular Prosthetic Limb (MPL, Johns Hopkins, Applied Physics Laboratory) (Johannes et al., 2011; Johannes et al., 2020). The motion of the robot hand was confined to the frontal plane and the subject controlled the horizontal and vertical velocity. The horizontal position of the robot hand was used to identify epochs corresponding to leftto-right and right-to-left reaches. Each reach was identified as the segment in which the robotic arm started from one local positional extrema (left or right), and moved to the opposite positional extrema (right or left). In the conditions where the participant was receiving no sensory feedback or incongruent sensory feedback, segmenting by the position of the robot arm resulted in the inclusion of segments where the participant did not successfully cross the target lines. We therefore identified each reach epoch as the segment between one positional extrema to the time point corresponding to the median length across all trials. Neural firing rates were smoothed with a 180 ms Gaussian kernel and segmented into the same reach epochs.

### Principal components analysis within sensory feedback conditions

During the task, the BCI user attempts to control the neural population activity to complete the task effectively, but the pattern of neural activity may also depend on the presence of visual and/or proprioceptive feedback. Typically, BCI control tasks are performed only with visual feedback. However, as M1 contains neurons that receive proprioceptive inputs, adding proprioceptive feedback may alter the firing rates of these neurons, thus shifting the neural population activity vector and the velocity control outputs from the decoder. Since N> > 2, the decoder represents one of many possible mappings from the N-dimensional space of the neurons to the 2D plane of the decoder output.

Training the decoder with vision alone results in a mapping that is optimized for that condition, but an alternate mapping may likely work better when proproprioceptive feedback is added. To explore alternate mappings, we used principal components analysis (PCA) to find axes of neural activity that captured variance within each sensory feedback condition. Neural activity segmented into left and right reaches was used to identify the axes that best explained the variance in neural activity for each sensory feedback condition. To ensure that movement direction contributed equal variance to the training data, trials for each sensory feedback modality were subsampled so that each crossing direction was equally represented in the training data. Subsampling in this way accounts for crossings that vary in length and prevents longer crossings from dominating the variance of the overall neural activity used for PCA. This was done ten times with non-overlapping training sets each time to provide a 10-fold cross-validated estimate of the PC space that captured the most variance in neural activity for each condition.

To quantify how proprioceptive feedback during the line-crossing task influenced the BCI decoders trained with vision only or with both vision and proprioception, we compared how well decoders captured neural variance across different sensory feedback conditions. To do so, we represented the neural subspace by taking the top *n* principal components that explained at least 90% of the variance, and computed the principal angle between this space and the horizontal decode axis. A smaller angle would mean that there is greater overlap between neural activity and the decode axis, meaning that the fluctuations in neural activity contribute more to fluctuations along the decode axis and therefore the decoded velocity. In contrast, an angle closer to 900 suggests that the projection of neural activity onto the decode axis is smaller, so variance in neural activity has less influence on the decoded velocity.

### Neural gradient analysis

We next determined whether sensory feedback resulted in smoother dynamics, or more consistent time evolution of neural activity. To do so, we took the top *n* PC components that explained 90% of the variance of the neural population activity across all conditions and divided this space into a number of bins such that each bin explained 5% of the variance of the overall neural activity. Since each PC dimension explains a different amount of variance of neural activity, each dimension is divided into a different number of bins. This allows dimensions that explain more of the overall variance to be divided into more bins, while PCs that explain less of the overall neural variance are divided into fewer bins, ensuring that each bin in neural space captures the same amount of variance of the overall neural activity. For example, if the first PC explained 30% of the overall variance, it would be divided into 6 bins, and if the second PC explained 10% of the variance, it would be divided into 2 bins. Together, this discretization of the PC space would result in a space comprising of 12 bins. The gray lines in Figure 7B illustrate how the PC space was divided into bins.

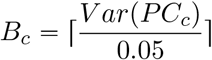

*B*_*c*_ is the number of bins for principal component *c*, and *V ar* (*PC*_*c*_) is the variance of the overall neural activity explained by principal component *c*. To completely discretize the PC space into bins, the range of PC values for a single component *c* is divided into the number of bins computed above, *B*_*c*_. This gives a set of scalars of size *B*_*c*_ for each component, where each value denotes a bin edge along PC axis *c*. Each of these bin edges is then paired with all other bin edges for all other components to create a set of *C* dimensional vectors, where *C* is the number of components that explains 90% of the overall neural variance. Each bin is therefore delineated by two *C* dimensional vectors, where any neural activity that falls between the edges of each bin is considered as a point in neural space within that bin. This allows us to divide the neural subspace into a set of discrete states. For each bin, we identify all points at which neural activity falls within that bin and compute the derivative of the neural activity with respect to time. This results in a set of vectors for each bin that represent the time derivative of the neural firing rates projected into the space of the *n* principal components. We normalize all vectors in a given bin to focus on the direction of the neural trajectories and not the speed of the neural trajectory. To estimate the consistency of the neural trajectory at any bin in the PC space, we average all the neural derivative vectors in a single bin. The length of the mean gradient vector will therefore be longer if the collection of vectors are pointing in consistent directions and shorter if the collection of vectors are pointing in disparate directions. Figure 7B shows an example of the derivative of the neural activity in two bins for two hypothetical trajectories, indicated by the blue or red traces. In each bin, these derivative vectors are normalized and the length of the average vector represents a measure of the consistency of the neural activity over time for any given point in the neural subspace. If the evolution of neural activity at a given point in neural space is consistent, then the average vector length will be longer (Figure 7C, top) than if the vector orientation is highly variable over time (Figure 7C, bottom).

## Results

The goal of this study was to examine how proprioceptive and visual feedback influence neural activity in M1 during BCI control of reaching with a robotic arm. Providing a BCI user with proprioceptive feedback enabled her to successfully control a robotic arm. However, proprioceptive feedback also influenced behavior due to interactions with M1 activity.

### Visual or proprioceptive feedback is required to perform the line-crossing task

Task performance was measured by counting the number of times the robot arm crossed the target lines (Figure 2). Performance on the task differed across sensory feedback conditions and interactions between decoder training and sensory feedback conditions, but not between training conditions alone (multi-way ANOVA *F*_*decoder*_ 0.35, *decoder* = 0.55; *F*_*feedback*_ =103.88, *p*_*feedback*_ = 2.02 * 10^−46^; *F*_*interaction*_ = 5.51, *p*_*interaction*_ = 3.28 *10^−4^). The average number of crossings for each sensory feedback condition on each experiment day are shown in Figure 2A. The participant was able to perform the task successfully during all conditions that were tested, so long as the sensory feedback being provided was congruent; without visual or proprioceptive feedback, performance was very poor (Figure 2A, gray), as was performance when proprioceptive feedback was incongruent with movement (Figure 2A, orange).

**Figure 2:**
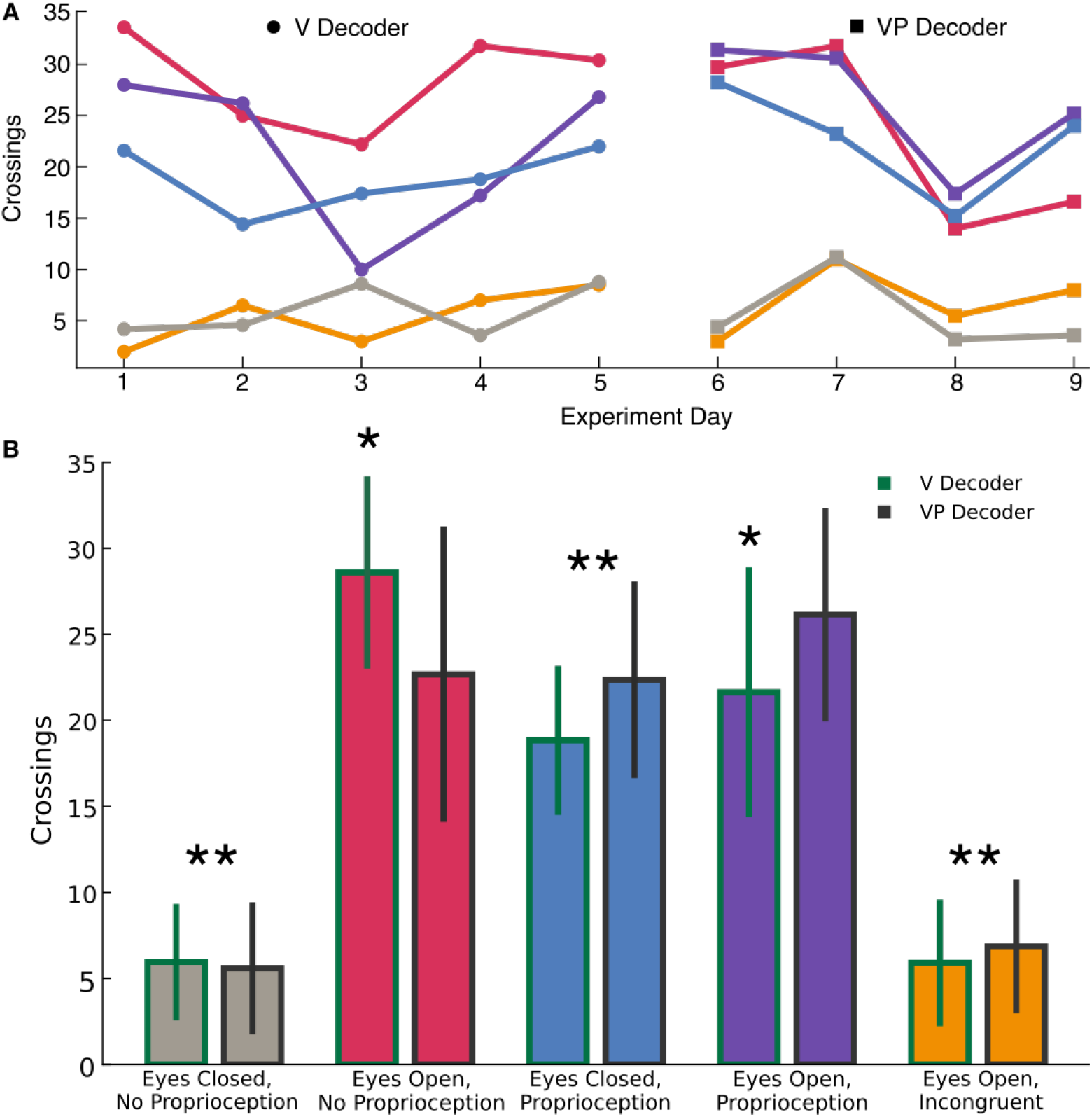
Summary of BCI Performance under various feedback conditions. A: Number of crossings on each day of the experiment. Color corresponds to sensory feedback condition during the BCI task. B: Number of crossings (mean and standard deviation across days) all combinations of sensory feedback. Green outline (left) is average of all days with the V Decoder, and black outlines (right) indicate average of all days with the VP decoder. * = the Eyes Open, No Proprioception condition with the V decoder is better than the Eyes Open, Proprioception condition for the same decoder; ** = the Eyes Closed, Proprioception condition is better than both the no feedback and the incongruent proprioceptive feedback trials.

Average crossings for each sensory feedback condition are shown in Figure 2B. Providing proprioceptive feedback alone recovered performance compared to when no feedback was provided (*p <* 1 * 10^−10^, ** in Figure 2B), and the same was true when visual feedback alone was provided (*p <* 1*10^−10^), for both decoders. However, incongruent proprioceptive feedback inhibited performance as much as no sensory feedback (Figure 2B, *p* > 0.95 for both decoders).

### Proprioceptive feedback impairs performance unless decoders are trained with proprioception

Although proprioceptive feedback rescued performance compared to no sensory feedback for the V decoder, performance on the task was more varied compared to when the participant was provided with visual and proprioceptive feedback. In contrast, training the decoder with vision and proprioception made BCI performance on these days (days 6-9) more robust to the addition of proprioceptive feedback during the task (Figure 2A, purple trace is consistently comparable to blue and pink traces for VP decoder but not V decoder). Averaged across days, providing proprioceptive feedback impaired performance when a decoder was trained with vision alone, as measured by the average number of reaches (line crossings) across all days (*p* = 1.3 * 10^−3^, Figure 2B, indicated by *). However, when the decoder was trained with both vision and proprioception, providing proprioceptive feedback in conjunction with vision did not impair task performance (*p* = 0.6).

### Sensory feedback conditions influence kinematics

Providing visual or proprioceptive feedback differently influenced the kinematics of the robotic arm, suggesting that sensory feedback influenced neural activity in M1 and through this, decoder performance. Figure 3A shows the horizontal position of the robot hand during representative trials when the decoder was trained with both vision and proprioception. When performing the task without sensory feedback, the robotic arm drifted to the right side of the workspace, but the hand continued to alternate leftward and rightward throughout the trial. When either visual or proprioceptive feedback was provided, the participant was able to move the robotic arm symmetrically in both directions. The velocity vs. position traces for individual trials of each sensory feedback condition on day 6 of the experiment (VP Decoder) are shown in Figure 3B.

**Figure 3:**
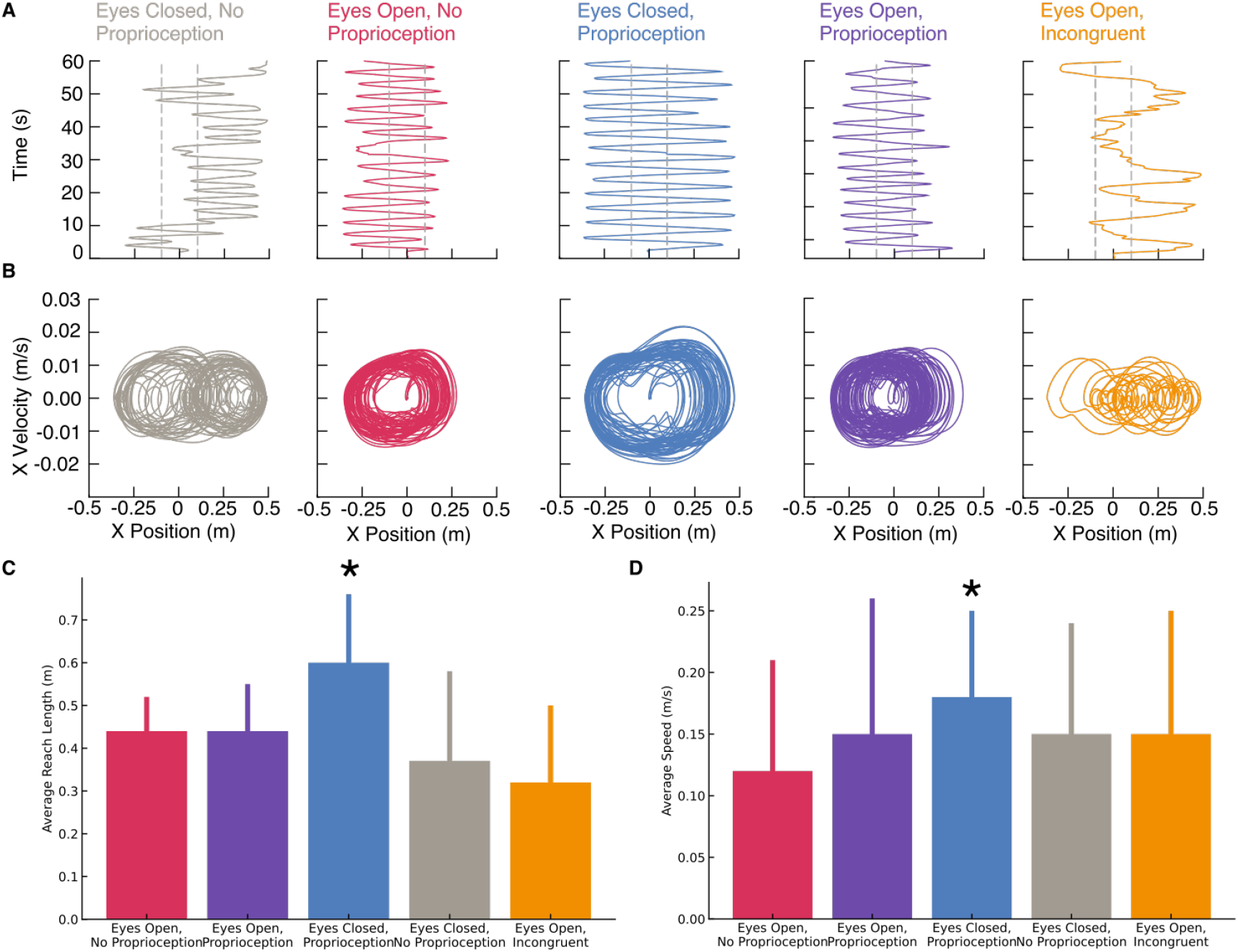
Kinematics of BCI control under various feedback conditions. A and B: Sample kinematics of the robotic arm on a VP decoder day. Time vs. horizontal position of robot arm (A) or horizontal position vs. horizontal velocity (B). Dashed gray lines correspond to markings the participant was asked to move robot arm across. Reach/crossing length (C) and (D) speed averaged across decoder types for each sensory feedback condition

When visual feedback was provided, regardless of whether proprioceptive feedback was provided, the BCI-controlled movements were more accurate, with each reach passing just over the right and left boundaries before changing direction. In contrast, in trials without visual feedback, the addition of proprioceptive feedback resulted in longer reaches and movements that were faster. Reach lengths were longer when proprioceptive feedback was the only modality provided compared to any of the other visual feedback conditions (ANOVA with Tukey post-hoc comparison (*F* = 33.9, *p* = 1.59 * 10^−20^, Eyes Open, No Proprioception: *p=* 0.0; Eyes Open, Proprioception: *p=* 2.26 * 10^−13^; Eyes Open, Incongruent: *p* = 2.50 *10^−10^, Figure 3C). Similarly, movements when proprioceptive feedback was provided in the absence of vision were faster than movements when visual feedback was provided (*p <*0.049, Figure 3D).

Since reach length and speed were higher when proprioception was the only available feedback modality, but similar when vision was available, regardless of whether proprioception was provided, it is likely that the difference in kinematics was due to a strategy change on the part of the participant as opposed to a proprioceptive effect on M1 activity. If the increase in speed in the Eyes Closed, Proprioception condition had been due to an effect of proprioception on M1 activity, then we would have expected to see increases in movement speed across all conditions in which proprioceptive feedback is provided.

Combined, these results raise the question of how proprioceptive feedback interacts with the control of movement in M1 to impair or improve performance. To address this question, we first examined how proprioceptive feedback influenced the population activity of neurons in M1. Then we examined how training a BCI decoder with visual feedback or combined visual and proprioceptive feedback influences population neural activity driven by different sensory feedback conditions.

### Proprioceptive feedback decreases M1 activity when provided in combination with visual feedback

Previously, proprioceptive feedback during BCI control has been shown to increase the firing rate of neurons in M1 (Suminski et al., 2010). We therefore examined the average firing rates of each channel across different sensory feedback conditions during the task. For each channel, we averaged the firing rate across all trials of a given sensory feedback condition and plotted it against the average firing rate in another sensory feedback condition (Figure 4). Points that fall along the dashed gray line indicate neurons that have the same average firing rate across the two sensory feedback conditions indicated on the horizontal and vertical axis labels. The green and black lines respectively show the linear regression model fit between the average firing rates across the two sensory feedback conditions for channels used in the V decoder and the VP decoder, respectively. A linear model with a slope below the dashed gray lines indicate that neurons on average had lower activity in the sensory feedback condition indicated by the vertical axis than in the sensory feedback condition indicated by the horizontal axis. In contrast to previous studies (Suminski et al., 2010), we found that firing rates of individual channels decreased when proprioceptive feedback was provided in combination with visual feedback (Figure 4A; t-test with Bonferroni correction *p*_*V*_ < 0.005; *p*_*V P*_ < 0.005), but increased when proprioceptive feedback was provided without visual feedback (Figure 4B;*p*_*V*_ < 0.05; *p*_*V P*_ < 0.005). However, firing rates for the Eyes Open, No Proprioception condition were similar relative to the Eyes Closed, Proprioception condition (Figure 4C; *p* > 0.05 for both decoders). Based on the changes in average firing rates when proprioception is provided or not provided across the Eyes Open and Eyes Closed conditions, the influence of proprioception on M1 activity depends on the presence of visual feedback. The differences in average firing rates between the visual-only or proprioception-only conditions between the two decoder types also suggests that there are interactions between motor activity during BCI control and sensory feedback that influences neural activity in M1. Thus, the influence of proprioceptive feedback on M1 activity depends not only upon the presence of visual feedback during the task, but also the sensory feedback provided during decoder training.

**Figure 4:**
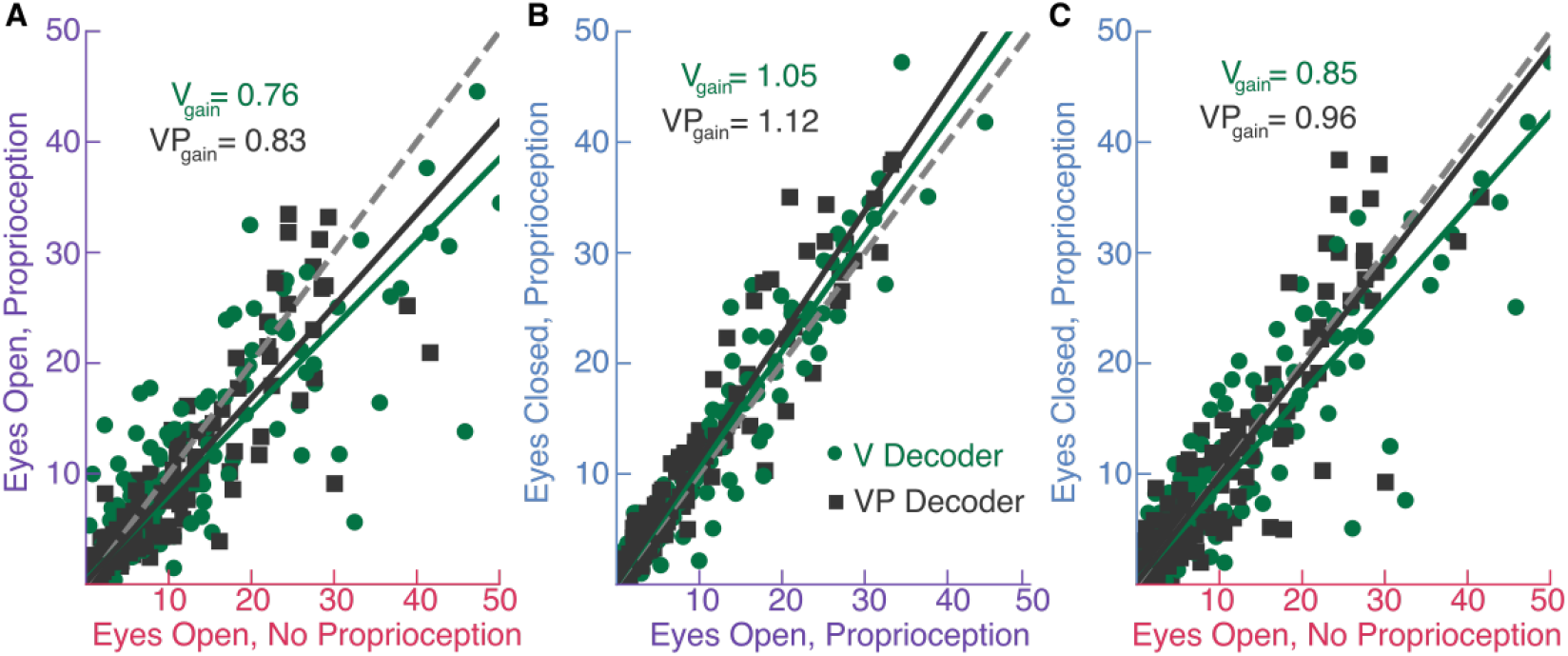
Changes in firing rates of M1 neurons across sensory feedback conditions. A: Average firing rate for each channel in the Eyes Open, No Proprioception condition is plotted against the average firing rate for each channel in trials with the Eyes Open, Proprioception condition. Green circles indicate a channel on a day when the decoder was trained with vision only, a black square indicates a channel on a day when the decoder was trained with vision and proprioception. B: Same as (A) but the average firing rate in the Eyes Open, Proprioception condition is plotted against the Eyes Closed, Proprioception condition. C: Same as (A) and (B) for the Eyes Open, No Proprioception condition and the Eyes Closed, Proprioception condition. Dashed gray line is the unity line. Points that fall on this line are units that fire equally in both sensory feedback conditions. Solid lines are the linear best fit for the relationship between average firing rates for the two sensory feedback conditions for the V decoder (green) and the VP decoder (black). Gains (slopes) for each line are labeled.

### Training the decoder with vision and proprioception allows the decoder to take advantage of proprioceptive driven signals in M1

To better understand how changes in the firing rates of individual neurons affected the overall population activity, we next examined how providing proprioceptive feedback during decoder training interacted with proprioceptive feedback during BCI control. Using PCA, we identified the axes that best captured the variance of neural activity in each sensory feedback condition on each experiment day. Sample neural trajectories for each sensory feedback and decoder condition are shown in Figure 5A-B. Trajectories in the green rectangle are trajectories where the V decoder was used (Figure 5A), and those in the gray rectangle are trajectories where the VP decoder was used (Figure 5B). The 2-headed arrow indicates the decode axis projected into each PC space. When the V decoder is used but proprioceptive feedback is provided with visual feedback during the task, neural trajectories are less smooth compared to when task feedback matches decoder feedback (Figure 5A, right).

**Figure 5:**
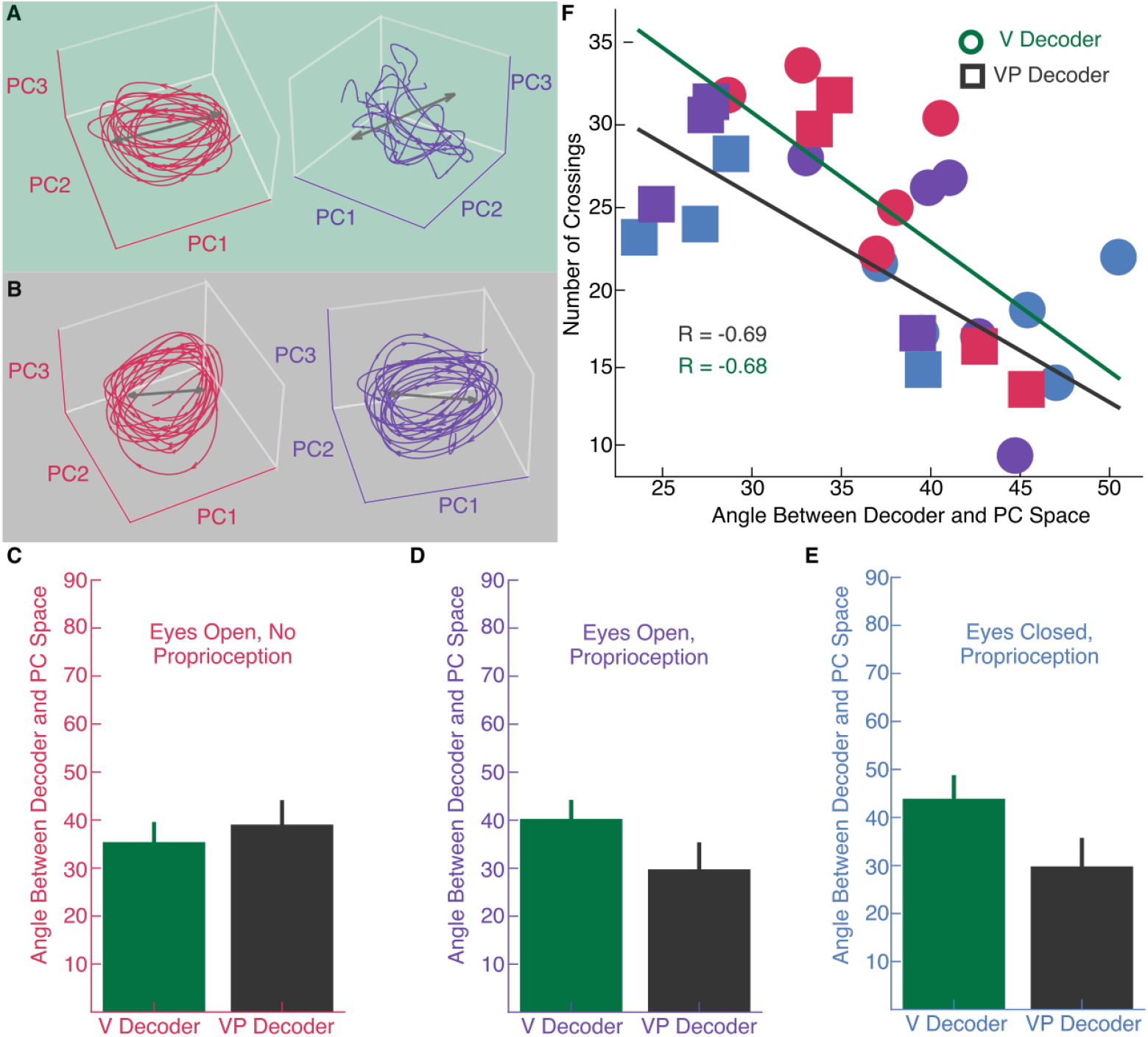
Neural subspace analysis across sensory feedback conditions. A: Example neural trajectories in the top 3 PCs for the Eyes Open, No Proprioception condition (left, red traces) and the top 3 PCs for the Eyes Open, Proprioception condition (right, purple traces) on a day when the decoder was trained with vision alone. B: Example neural trajectories for the same conditions as in (A) on a day when the decoder was trained with vision and proprioception. C: The angle between the neural subspace defined by the set of basis vectors identified via PCA and the decode axis averaged across all days for each decoder for the Eyes Open, No Proprioception condition D: Same as (C) for the Eyes Open, Proprioception condition E: Same as (C, D) for the Eyes Closed, Proprioception condition. Error bars indicate standard deviation across days. F: Average number of line crossings plotted against the angle between decode axis and PC axis for each sensory feedback condition on each day. Circular markers indicate days in which the decoder was trained with vision only, square markers indicate days in which the decoder was trained with vision and proprioception. The number in each marker indicates the day in the overall experiment. Solid lines indicate lines of best fit between the principal angle and the number of reaches for the V Decoder (green) and VP decoder (black)

We then measured the principal angle between this low-dimensional neural subspace that encompasses 90% of the overall variance of neural activity for each sensory feedback condition and the horizontal decode axis. The principal angle estimates the projection length of the neural subspace defined by sensory feedback on the decode axis. An angle of 90 ° means that there is the projection length between the neural variability driven by sensory feedback and the decode axis is zero, and therefore the variation in neural activity due to sensory feedback during the task does not influence neural activity along the decode axis. Conversely, a principal angle of 0^*circ*^ indicates that sensory feedback drives variability in neural responses along the same axis as the decoder. We found that when proprioceptive feedback was provided during the task, variation in neural activity aligned better with the decode axis, because the angle between the PC space capturing 90% of neural variance and the decode axis was smaller for the vision and proprioception decoder than the vision only decoder (Figure 5D, E). In contrast, when proprioceptive feedback is not provided, the angle between the decode axis and neural activity is not different across decoders (Figure 5C). Thus, training the decoder with vision and proprioception ensures that proprioceptive-driven features of M1 neural population activity are taken into account by the decoder, as the principal angle between the decode axis and proprioceptive-driven variability in neural activity is smaller.

Across all conditions where the participant was receiving sensory feedback, the number of crossings on each day was negatively correlated with the overlap in the PC space and the decoder axis (V Decoder = *R =* −0.68, VP Decoder = *R =* −0.69) (Figure 5F). Thus, performance is better when neural activity during the task aligns well with the decoder axis.

To ensure that this alignment between neural variability and the decoder axis was due to decoder training and not to the participant’s ability to successfully use the BCI, we examined the relationship between neural activity in the incongruent proprioceptive feedback condition and the decode axis. We found that the angle between the decoder and neural activity during BCI control was smaller for the VP decoder

(37.47° ±5.46) than the V decoder (61.45° ±7.92), indicating that training the decoder with proprioceptive feedback better encapsulates the proprioceptive space (Figure 6A). However, despite good alignment between neural activity and the decode axis, task performance in the eyes open, incongruent condition was poor, and there was no correlation between the overlap between the decoder and neural activity during BCI control and BCI performance (*R =* −0.24, Figure 6B). To summarize, training a decoder with proprioception allows the decoder to capture proprioceptive-driven signals, and erroneous proprioception disrupts performance.

**Figure 6:**
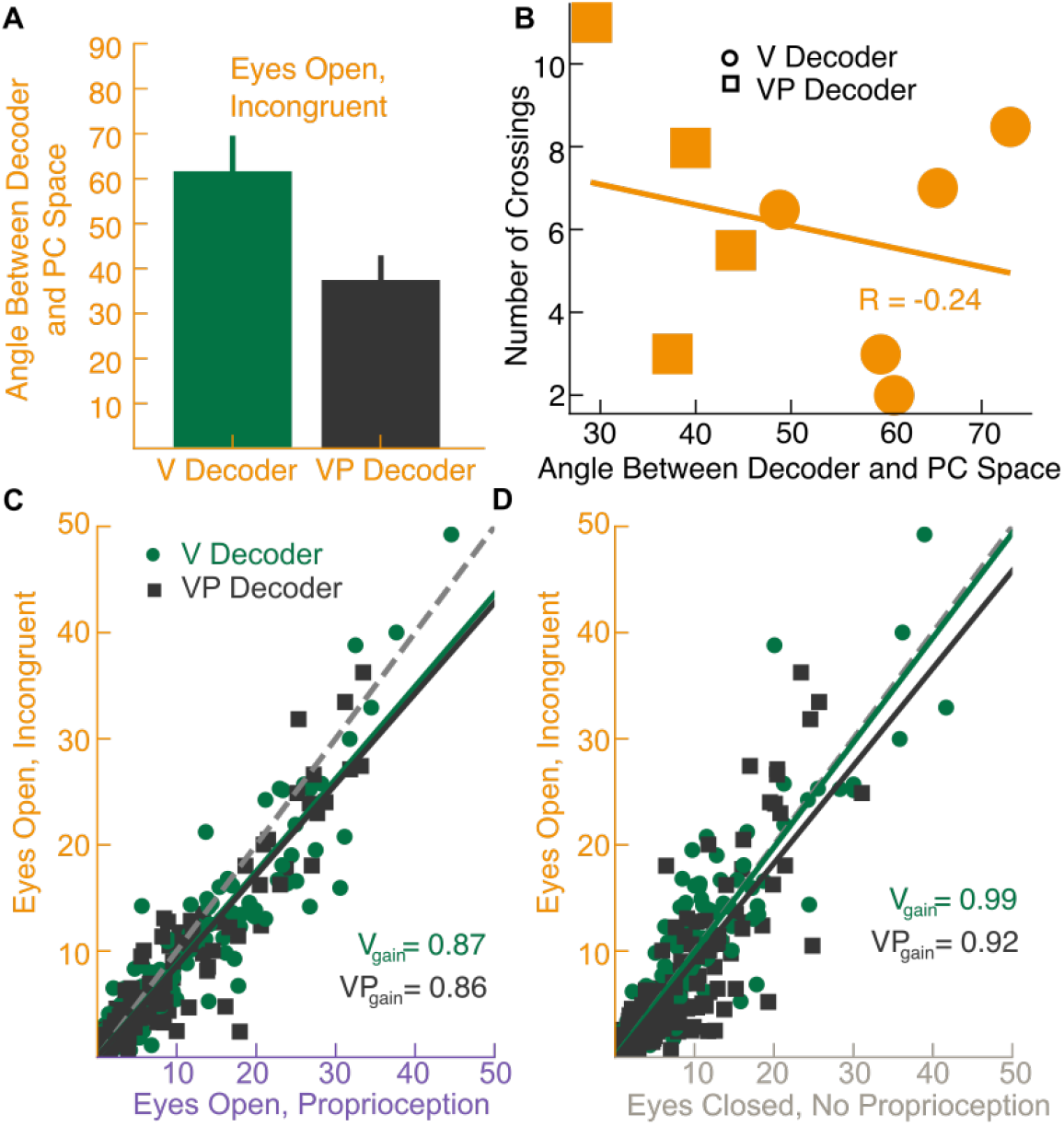
Summary of neural analysis of incongruent proprioception condition. A: Angle between the PC space and the decode axis averaged across all days for each decoder for the eyes open, incongruent condition. B: Average number of crossings for the eyes open, incongruent condition on each day plotted against the angle between decode axis and PC space. C: Average firing rate for each channel in trials with the eyes open, incongruent proprioception condition is plotted against the average firing rate for each channel in the eyes open, proprioception condition. D: Same as (C) but the average firing rate in the eyes open, incongruent proprioception condition is plotted against the eyes closed, no proprioception condition.

To understand how incongruent proprioception influences neural activity in M1, we compared the average firing rates of M1 neurons in the incongruent proprioception condition to the eyes open, proprioception condition. Interestingly, we found that across both decoder types, firing rates across neurons in M1 were reduced relative to the eyes open, proprioception condition (Figure 6C; t-test with Bonferroni correction *p* < 0.005 for both decoders), suggesting that some activity in M1 is inhibited due to erroneous sensory feedback. Additionally, firing rates were slightly reduced compared to the no sensory feedback condition for the VP decoder (*p* < 0.005) but not the V decoder (*p* > 0.05), further reinforcing the finding that sensory feedback provided during decoder training influences how sensory feedback during the task modulates M1 activity (Figure 6D). To better understand the influence of sensory feedback on BCI control, we next examined how neural activity evolved over time in a single trial.

### Smooth neural trajectories correlate with good control

After establishing that overlap between the decode axis and neural activity in each sensory feedback condition was correlated with task performance, we wanted to determine how sensory feedback influenced the time-varying neural activity over the course of a single trial. To do so, we examined how neural activity evolves over time in the PC space that explains at least 90% of the variance of neural activity in each sensory feedback condition. Example neural trajectories for single trials are shown in Figure 5A (V Decoder) and 5B (VP Decoder). When the decoder was trained with vision alone (V Decoder, Figure 5A), neural trajectories were smooth for the eyes open, no proprioception condition, but did not display smooth trajectories in the eyes open, proprioception condition. In contrast, when the decoder was trained with vision and proprioception (VP Decoder, Figure 5B), neural trajectories were smooth across both conditions. Thus, sensory feedback influences how smoothly neural trajectories evolve over the course of a single trial.

To quantify the smoothness of the neural trajectories for each sensory feedback condition, we divided the PC space for the top n components that explains 90% of the variance into small bins (Figure 7B,C). We graded bin sizes so that axes that explained more variance were divided into smaller bins and axes that explained less variance were divided into larger bins. At each time point during which neural activity passed through a particular bin, we computed the derivative of the neural activity with respect to time. This allowed us to visualize the direction in which neural activity was moving in time at each point in PC space by providing a single vector for each time point. For each point in PC space, we then averaged these vectors of neural activity to estimate a flow field of neural activity for each condition (Figure 7A). This is similar to the tangling metric introduced in Russo et al., 2018, but allows us to better capture points in neural space where we have few samples via discretization.

**Figure 7:**
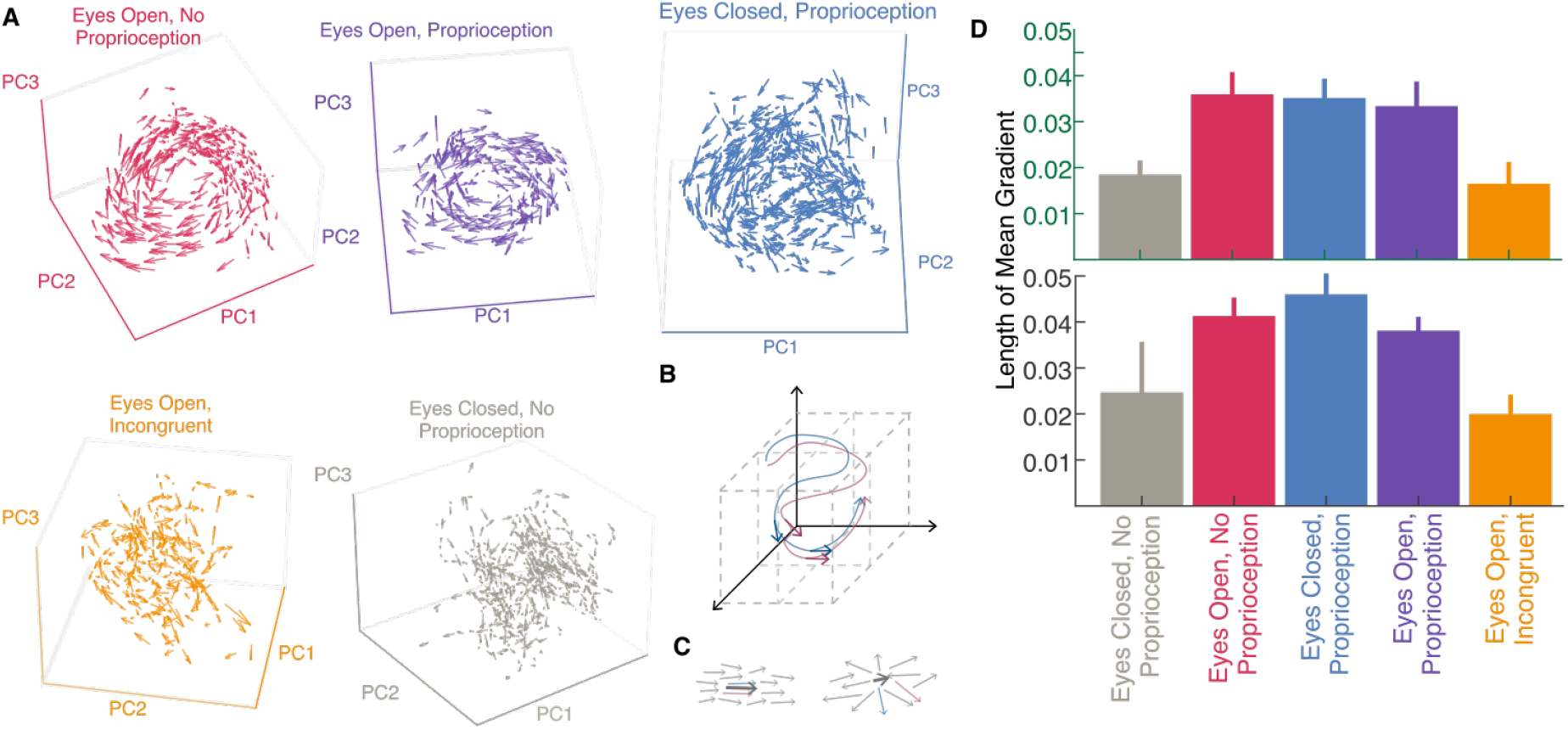
Smoothness of neural trajectories during BCI control. A: Average neural gradients visualized at each point in the top 3 PCs in five different feedback conditions. B: The PC subspace explaining 90% of the variance was divided into n dimensional bins, with 5% of the total variance explained per bin. The time derivative of the neural trajectory passing through each bin was calculated to build a set of vectors in each bin. These vectors were normalized and averaged to derive a metric of how consistent neural dynamics were at different neural states. C: When vectors in a particular bin have a very consistent direction as in the top example, the mean vector (dark black arrow) will be longer in length than when vectors in a bin do not point in similar directions (bottom example). D: Average length of the mean gradient vector across all bins in neural space for each sensory feedback condition for a vision only decoder (top) and a vision and proprioception decoder (bottom)

We estimated the smoothness of the neural trajectories by computing the length of the average gradient vector of the trajectories at different points in neural space. If neural trajectories are smooth, then we would expect the length of the average vector to be long, because the gradients in each point are pointing in the same direction. In contrast, if the neural trajectories are not smooth, then we would expect the length of the average gradient vector to be short because disparate directions average out to a zero-length vector. We found that the mean normalized gradient length was longer when visual and proprioceptive feedback were provided alone or combined, than when no sensory feedback was provided or incongruent proprioceptive feedback was provided, suggesting that neural state is more predictive of where neural activity will go when sensory feedback is provided (Figure 7D) (ANOVA with *post hoc* t-test; *p* < 0.005 for all sensory congruent sensory feedback conditions). In contrast, when no sensory feedback or incongruent proprioceptive feedback was provided, neural gradients were less smooth and more variable. A decoder trained with vision and proprioception, had longer gradients than a decoder trained with vision only, suggesting that volitional control of movement via a BCI may be influenced by sensory feedback provided during decoder training. Together, these results suggest that when sensory feedback is provided, neural trajectories evolve more smoothly over time and results in smoother control of the robotic hand.

## Discussion

Proprioceptive feedback is crucial across many stages of motor control. We found that during BCI control, M1 responds to proprioceptive feedback in complex ways that depend on both the presence of visual feedback and the sensory modalities provided while training the BCI decoder. Understanding these interactions can offer insight into both sensory restoration for BCI users as well as decoder training paradigms that take into account multiple modalities of sensory feedback.

### Proprioceptive feedback impairs performance when decoder is trained with vision only

We observed that providing proprioceptive feedback during the task (eyes open, proprioception condition) impaired performance relative to providing visual feedback alone (eyes open, no proprioception), but only when the decoder was trained on visual feedback only (V Decoder). This contrasts with a previous study by Suminski et al., 2010, which found that when congruent visual and proprioceptive feedback were provided, performance in a random target pursuit task improved. Several differences in experimental design may explain these differences in results. First, the decoder in Suminski et al., 2010 was trained to generate a position signal from neural activity, whereas the decoder used here generated velocity commands from neural signals. Since previous studies have shown that proprioceptive feedback is essential for generating motor commands for movement velocity and joint torques (Sober & Sabes, 2003), it is possible that proprioceptive feedback has a greater effect on velocity decoders than position decoders. Taken together, these findings raise interesting questions as to how M1 integrates proprioceptive feedback—if differences between Suminski et al., 2010 and our results are primarily due to the decoded kinematic parameter (position or velocity), this suggests that proprioceptive feedback in M1 is largely informative about arm velocity, but that M1 can transform velocity feedback into a positional command. Further experiments are needed to examine the neural coding of velocity or position in M1, as well as a BCI user’s ability to transform velocity information into position commands and vice versa.

### Training a decoder with visual and proprioceptive feedback restores performance

While proprioceptive feedback paired with a V decoder impaired performance, training the decoder with visual and proprioceptive feedback restored performance to that of a V decoder paired with visual only feedback. One explanation for this is that training the decoder with vision and proprioception increased the overlap between neural activity and the decode axis. In other words, proprioceptive feedback influences M1 activity, and a V decoder is not guaranteed to fit a decode axis that is well-aligned with the proprioception-driven variability. Training a decoder with vision and proprioception, however, ensures there is good alignment between the decode axis and the proprioception-driven variability in M1 activity. In some regards, this result is intuitive in that the proprioceptive feedback is well-correlated with velocity during decoder training, and therefore a straightforward linear estimation will capture the variance resulting from proprioceptive-modulated neurons in M1. However, we would like to note that if this were the case, we might expect to see that performance in the incongruent proprioceptive feedback condition could be as good as in the proprioception-only condition if the participant simply lets the proprioceptive signal dominate control. In contrast, we see that incongruent proprioceptive feedback significantly impairs performance, suggesting that proprioceptive feedback is not simply driving neural activity along the decode axis, but that proprioceptive feedback interacts with the participant’s volitional control of the BCI in some way.

### Proprioception directly influences M1 activity

The finding that proprioceptive feedback influences BCI performance is unsurprising given that M1 has been shown to respond to proprioception and other forms of sensory feedback (Evarts & Fromm, 1977; Hatsopoulos & Suminski, 2011). In accordance with many of these studies, we found that proprioceptive responses in M1 vary depending on whether the participant’s eyes were open or closed. More specifically, proprioceptive neurons appeared to increase firing in the absence of visual feedback and decrease firing in the presence of visual feedback. Such differential activation can be the result of several possibilities: 1) the participant is able to, via gating or other methods, volitionally “turn off” proprioceptive neurons when visual feedback is provided, because visual feedback is more reliable for movement direction and arm positioning (Rosenkranz & Rothwell, 2012; Seki & Fetz, 2012); 2) M1 neurons encode errors in sensory estimation, and when congruent visual and proprioceptive feedback are provided, error cancellation between visual and proprioceptive feedback reduces firing (Inoue et al., 2016); and 3) sensory state representations are integrated across sensory feedback modalities to generate a motor command, and when congruent visual and proprioceptive feedback are provided, the participant generates a smoother movement and therefore a less noisy motor plan (Wolpert & Ghahramani, 2000).

The decrease in firing rate due to proprioceptive feedback that we observed seems unlikely to be caused by a gating mechanism, as BCI performance is impaired in the eyes open, proprioception condition for the V decoder, suggesting that proprioception is influencing performance and its effect on M1 activity cannot be downmodulated. Additionally, the participant is unable to perform the task when incongruent proprioceptive feedback is provided, suggesting that erroneous proprioceptive feedback cannot be ignored. If the decrease in firing rate is due to some cancellation between visual and proprioceptive feedback, we would expect that firing rates are also higher for the eyes open, incongruent proproception condition relative to the eyes open, no proprioception condition. We did not find this to be the case, however, as firing rates were lower for the eyes open, incongruent proprioception condition compared to the eyes open, proprioception condition. Lastly, if smoother movements correspond to lower firing rates, then we would expect that the sensory feedback condition with best performance for each decoder type is also the sensory feedback condition with the lowest average firing rates. While this was true for the V decoder, it was not the case for the VP decoder, suggesting that there are interactions between decoder training and task feedback that contribute to how proprioceptive feedback influences M1 activity. Our findings are once again different from those of Suminski et al., 2010, but this could be due to differences in how M1 responds to proprioceptive feedback when the BCI user is generating position or velocity signals.

Further analysis into how M1 encodes movement and responds to sensory feedback based on velocity or position coordinates is needed.

We lastly examined the relationship between neural activity and kinematic control. Many recent studies have examined dynamics in M1, and the role of sensory feedback in these dynamics (Churchland et al., 2012; Lara et al., 2018; Russo et al., 2018; Shenoy et al., 2013; Suresh et al., 2020). We found that neural trajectories appeared smooth when control was good, and similarly, neural trajectories were not smooth when control was poor. If smooth dynamical trajectories are indicative of more innately dynamical systems, then external inputs such as sensory feedback would degenerate deviations from the dynamic behavior (Russo et al., 2018). In contrast, we found that sensory feedback made neural activity in M1 more dynamical. While we did not see smooth dynamical trajectories when no sensory feedback was provided, there are a number of reasons why this might be the case. One possibility is that sensory feedback is required to drive smooth dynamics in M1—particularly in the context of a task. Another possibility is that BCI control is very different from natural arm control. In particular, in many of the tasks which examine dynamics, monkeys are using natural arm movements in reaching tasks (Churchland et al., 2012; Lara et al., 2018; Russo et al., 2018). Thus, it may be that when the monkey is using natural arm movements that they are already familiar with, dynamical activity in M1 is smooth and can help drive motor control and provide signals to downstream motor neurons. In contrast, control of a BCI decoder may utilize a different mechanism and require more sensory feedback to drive control in M1.

### When visual feedback is not provided, proprioceptive feedback provides useful information

Although proprioceptive feedback impaired performance when provided alongside visual feedback to a V Decoder, this feedback was still sufficient for the participant to perform the task successfully compared to having no feedback at all about arm position. However, different sensory modalities can be more or less informative about movement and therefore influence movement accuracy and precision. Indeed, we found that proprioceptive feedback in the absence of visual feedback led to larger amplitude movements at higher speed. Since this increase in velocity occurred only in the eyes closed, proprioception condition but not in the eyes open, proprioception condition, it is likely that this effect is due to a shift in participant strategy rather than a result of proprioceptive feedback on M1, or that visual feedback is slower. Previous studies have shown that removal of visual feedback during reaching tasks induces accumulating drift in arm position from the optimal trajectory (Desmurget et al., 1997; Ghez et al., 1995), although the direction and distance of movement remains relatively constant (Brown et al., 2003). The strategy shift we saw is in line with these studies in that without a visual cue to guide robot arm position, an effective way to maximize line crossings is to move quickly over longer distances. From these results, one might expect that when no sensory feedback was provided, the participant might adopt a strategy similar to her strategy in the proprioceptive feedback-only condition. However, when no sensory feedback is provided, the robotic arm drifts to one side and continues to make leftward and rightward movements that are smaller in amplitude and slower. Thus, in accordance with previous literature that shows that proprioceptive feedback is essential for generating motor commands and movement extent (Brown et al., 2003; Scheidt et al., 2005; Sober & Sabes, 2003), the movements in the no sensory feedback condition were neither positionally centered nor large in amplitude.

### Implications for BCI control

One of the major goals of examining the impact of sensory feedback in M1 is evaluating the potential benefits of sensory restoration for BCI users. Sensory restoration via intracortical microstimulation (ICMS) (Fifer et al., 2022; Flesher et al., 2016; Flesher et al., 2021; Hughes et al., 2021; Shelchkova et al., 2023) has been shown to provide valuable feedback to BCI users, although ICMS can impair BCI control, as evoked activity in M1 due to ICMS disrupts the mapping between neural activity and velocity (Shelchkova et al., 2023). One advantage of proprioceptive feedback, however, is that it allows the BCI user to make relatively accurate movement in the absence of visual feedback. This is perhaps not surprising in that proprioceptors encode limb position as well as movement direction and velocity (Bosco & Poppele, 2001; Weber et al., 2011) variables that definitionally change with movement. Thus, developing ways to provide proprioceptive feedback to BCI users will broaden the range of tasks and activities in which BCI control can be used.

The results presented here, when combined with results by Suminski et al., 2010 that show that providing proprioceptive feedback to a position decoder trained with visual feedback only, indicate that more works needs to be done both in understanding how different sensory feedback modalities impact neural activity in M1, as well as the coordinates by which M1 controls and represents movement. For example, one might imagine that M1 is highly sensitive to velocity signals of the proprioceptors, but that training the decoder to output position rather than velocity requires the BCI user to transform positional signals to velocity signals and therefore reduces the direct impact of velocity-based proprioceptive signals on M1 activity. Therefore, understanding how M1 represents or integrates proprioceptive feedback into velocity or position signals can offer greater insight as to how to deliver proprioceptive feedback during BCI control. The study presented here had some limitations. We had a single participant (n=1) with intact somatosensation. Additionally, while our participant received a diagnosis of spinocerebellar degeneration, it is unclear whether this is the true pathology. Finally, proprioceptive feedback was provided via an experimenter moving the participant’s arm–more accurate proprioceptive feedback provided with lower latency would make the impacts of proprioceptive feedback on M1 more clear.

## Conclusion

We provided proprioceptive feedback to a person using a BCI decoder to perform a reaching task. We found that removing visual feedback significantly disrupted performance, but that providing proprioceptive feedback about the robotic arm recovered much of the task performance. Interestingly, however, when proprioceptive and visual feedback were provided together, task performance suffered if the decoder was trained with visual feedback alone. This penalty for including proprioceptive feedback was eliminated when the decoder was trained with both visual and proprioceptive feedback.This impairment seemed to arise because neurons in M1 respond to proprioceptive feedback in different ways depending on the presence of visual feedback and the sensory feedback provided during decoder training. Moreover, training the decoder with visual and proprioceptive feedback recovered decoder performance because the decoder was better able to account for the activity of proprioception-driven neurons in M1. Not only was the overlap between the decoder and the neural activity for each sensory feedback condition correlated with performance, the smoothness of the neural trajectory also correlated with the smoothness of the robot kinematics. Taken together with previous studies, these results suggest that proprioceptive feedback interacts with volitional control in M1 by impacting neural activity, which impacts decoder performance. Restoring sensory feedback to BCI may require taking into account the interactions between sensory feedback and volitional control in M1.

## Data Availability

All data produced in the present study are available upon reasonable request to the authors

## Acknowledgements

We thank Jan Scheuermann for her extraordinary commitment and effort related to this study as well as insightful discussions with the study team. We also thank Jeffrey Weiss for his engineering and data collection efforts.

This study was funded by the Defense Advanced Research Projects Agency’s (Arlington, VA, USA) Revolutionizing Prosthetics program (contract number N66001-10-C-4056). The views expressed herein are those of the authors and do not represent the official policy or position of the Department of Defense, or US Government.

## Notes

### Competing Interest Statement

The authors have declared no competing interest.

### Clinical Trial

This trial is registered on clinicaltrials.gov (http://clinicaltrials.gov/ct2/show/NCT01364480).

### Author Declarations

This study was conducted under an Investigational Device Exemption (IDE) granted by the US Food and Drug Administration and with approval from the Institutional Review Boards at the University of Pittsburgh and the Space and Naval Warfare Systems Center Pacific. This trial is registered on clinicaltrials.gov (http://clinicaltrials.gov/ct2/show/NCT01364480).

## References

Armenta Salas, M., Bashford, L., Kellis, S., Jafari, M., Jo, H., Kramer, D., Shanfield, K., Pejsa, K., Lee, B., Liu, C. Y., & Andersen, R. A. (2018). Proprioceptive and cutaneous sensations in humans elicited by intracortical microstimulation. eLife, 7, e32904. 10.7554/eLife.32904

Armiger, R. S., Tenore, F. V., Bishop, W. E., Beaty, J. D., Bridges, M. M., Burck, J. M., Vogelstein, R. J., & Harshbarger, S. D. (2011). A Real-Time Virtual Integration Environment. JOHNS HOPKINS APL TECHNICAL DIGEST, 30 (3), 9.

Boninger, M., Mitchell, G., Tyler-Kabara, E., Collinger, J., & Schwartz, A. B. (2013). Neuroprosthetic control and tetraplegia – Authors’reply [Publisher: Elsevier]. The Lancet, 381 (9881), 1900–1901. 10.1016/S0140-6736(13)61154-X

Bosco, G., & Poppele, R. E. (2001). Proprioception From a Spinocerebellar Perspective. Physiological Reviews, 81 (2), 539–568. 10.1152/physrev.2001.81.2.539 Publisher: American Physiological Society

Brown, L. E., Rosenbaum, D. A., & Sainburg, R. L. (2003). Limb Position Drift: Implications for Control of Posture and Movement. Journal of Neurophysiology, 90 (5), 3105–3118. 10.1152/jn.00013.2003 Publisher: American Physiological Society

Churchland, M. M., Cunningham, J. P., Kaufman, M. T., Foster, J. D., Nuyujukian, P., Ryu, S. I., & Shenoy, K. V. (2012). Neural population dynamics during reaching. Nature, 487 (7405), 51–56. 10.1038/nature11129

Collinger, J. L., Wodlinger, B., Downey, J. E., Wang, W., Tyler-Kabara, E. C., Weber, D. J., McMorland, A. J., Velliste, M., Boninger, M. L., & Schwartz, A. B. (2013). High-performance neuroprosthetic control by an individual with tetraplegia. The Lancet, 381 (9866), 557–564. 10.1016/S0140-6736(12)61816-9

Deo, D. R., Rezaii, P., Hochberg, L. R., Okamura, A. M., Shenoy, K. V., & Henderson, J. M. (2021). Effects of Peripheral Haptic Feedback on Intracortical Brain-Computer Interface Control and Associated Sensory Responses in Motor Cortex [Conference Name: IEEE Transactions on Haptics]. IEEE Transactions on Haptics, 14 (4), 762–775. 10.1109/TOH.2021.3072615

Desmurget, M., Jordan, M., Prablanc, C., & Jeannerod, M. (1997). Constrained and Unconstrained Movements Involve Different Control Strategies. Journal of Neurophysiology, 77 (3), 1644–1650. 10.1152/jn.1997.77.3.1644 Publisher: American Physiological Society

Evarts, E. V., & Tanji, J. (1976). Reflex and intended responses in motor cortex pyramidal tract neurons of monkey. Journal of Neurophysiology, 39 (5), 1069–1080. 10.1152/jn.1976.39.5.1069 Publisher: American Physiological Society

Evarts, E. V., & Fromm, C. (1977). Sensory responses in motor cortex neurons during precise motor control. Neuroscience Letters, 5 (5), 267–272. 10.1016/0304-3940(77)90077-5

Fetz, E. E., Finocchio, D. V., Baker, M. A., & Soso, M. J. (1980). Sensory and motor responses of precentral cortex cells during comparable passive and active joint movements. Journal of Neurophysiology, 43 (4), 1070–1089. 10.1152/jn.1980.43.4.1070

Fifer, M. S., McMullen, D. P., Osborn, L. E., Thomas, T. M., Christie, B., Nickl, R. W., Candrea, D. N., Pohlmeyer, E. A., Thompson, M. C., Anaya, M. A., Schellekens, W., Ramsey, N. F., Bensmaia, S. J., Anderson, W. S., Wester, B. A., Crone, N. E., Celnik, P. A., Cantarero, G. L., & Tenore, F. V. (2022). Intracortical Somatosensory Stimulation to Elicit Fingertip Sensations in an Individual With Spinal Cord Injury. Neurology, 98 (7), e679–e687. 10.1212/WNL.0000000000013173

Flesher, S. N., Collinger, J. L., Foldes, S. T., Weiss, J. M., Downey, J. E., Tyler-Kabara, E. C., Bensmaia, S. J., Schwartz, A. B., Boninger, M. L., & Gaunt, R. A. (2016). Intracortical microstimulation of human somatosensory cortex. Science Translational Medicine, 8 (361), 361ra141–361ra141. 10.1126/scitranslmed.aaf8083

Flesher, S. N., Downey, J. E., Weiss, J. M., Hughes, C. L., Herrera, A. J., Tyler-Kabara, E. C., Boninger, M. L., Collinger, J. L., & Gaunt, R. A. (2021). A brain-computer interface that evokes tactile sensations improves robotic arm control. Science, 372 (6544), 831–836. 10.1126/science.abd0380

Georgopoulos, A. P., Lurito, J. T., Petrides, M., Schwartz, A. B., & Massey, J. T. (1989). Mental rotation of the neuronal population vector. Science, 243 (4888), 234–236. 10.1126/science.2911737

Georgopoulos, A. P., Schwartz, A. B., & Kettner, R. E. (1986). Neuronal population coding of movement direction. Science (New York, N.Y.), 233 (4771), 1416–1419. 10.1126/science.3749885

Ghez, C., Gordon, J., & Ghilardi, M. F. (1995). Impairments of reaching movements in patients without proprioception. II. Effects of visual information on accuracy [Publisher: American Physiological Society]. Journal of Neurophysiology, 73 (1), 361–372. 10.1152/jn.1995.73.1.361

Hatsopoulos, N. G., & Suminski, A. J. (2011). Sensing with the Motor Cortex. Neuron, 72 (3), 477–487. 10.1016/j.neuron.2011.10.020

Hughes, C. L., Flesher, S. N., Weiss, J. M., Boninger, M. L., Collinger, J., & Gaunt, R. (2021). Perception of microstimulation frequency in human somatosensory cortex (J. A. Pruszynski, Ed.). eLife, 10, e65128. 10.7554/eLife.65128

Inoue, M., Uchimura, M., & Kitazawa, S. (2016). Error Signals in Motor Cortices Drive Adaptation in Reaching. Neuron, 90 (5), 1114–1126. 10.1016/j.neuron.2016.04.029

Johannes, M. S., Bigelow, J. D., Burck, J. M., Harshbarger, S. D., Kozlowski, M. V., & Doren, T. V. (2011). An Overview of the Developmental Process for the Modular Prosthetic Limb. JOHNS HOPKINS APL TECHNICAL DIGEST, 30 (3).

Johannes, M. S., Faulring, E. L., Katyal, K. D., Para, M. P., Helder, J. B., Makhlin, A., Moyer, T., Wahl, D., Solberg, J., Clark, S., Armiger, R. S., Lontz, T., Geberth, K., Moran, C. W., Wester, B. A., Van Doren, T., & Santos-Munne, J. J. (2020). Chapter 21 - The Modular Prosthetic Limb. In J. Rosen & P. W. Ferguson (Eds.), Wearable Robotics (pp. 393–444). Academic Press. 10.1016/B978-0-12-814659-0.00021-7

Lara, A. H., Elsayed, G. F., Zimnik, A. J., Cunningham, J. P., & Churchland, M. M. (2018). Conservation of preparatory neural events in monkey motor cortex regardless of how movement is initiated (J. L. Raymond & T. E. Behrens, Eds.). eLife, 7, e31826. 10.7554/eLife.31826 Publisher: eLife Sciences Publications, Ltd

Marquardt, D. W. (1970). Generalized Inverses, Ridge Regression, Biased Linear Estimation, and Nonlinear Estimation [Publisher: [Taylor & Francis, Ltd., American Statistical Association, American Society for Quality]]. Technometrics, 12 (3), 591–612. 10.2307/1267205

Moran, D. W., & Schwartz, A. B. (1999). Motor Cortical Representation of Speed and Direction During Reaching. Journal of Neurophysiology, 82 (5), 2676–2692. 10.1152/jn.1999.82.5.2676

Pellegrino, G. d., & Wise, S. P. (1993). Visuospatial versus visuomotor activity in the premotor and prefrontal cortex of a primate. Journal of Neuroscience, 13 (3), 1227–1243. 10.1523/JNEUROSCI.13-03-01227.1993

Pellizzer, G., Sargent, P., & Georgopoulos, A. P. (1995). Motor cortical activity in a context-recall task. Science, 269 (5224), 702–705. 10.1126/science.7624802

Rosenkranz, K., & Rothwell, J. C. (2012). Modulation of Proprioceptive Integration in the Motor Cortex Shapes Human Motor Learning. Journal of Neuroscience, 32 (26), 9000–9006. 10.1523/JNEUROSCI.0120-12.2012 Publisher: Society for Neuroscience Section: Articles

Russo, A. A., Bittner, S. R., Perkins, S. M., Seely, J. S., London, B. M., Lara, A. H., Miri, A., Marshall, N. J., Kohn, A., Jessell, T. M., Abbott, L. F., Cunningham, J. P., & Churchland, M. M. (2018). Motor Cortex Embeds Muscle-like Commands in an Untangled Population Response. Neuron, 97 (4), 953–966.e8. 10.1016/j.neuron.2018.01.004

Scheidt, R. A., Conditt, M. A., Secco, E. L., & Mussa-Ivaldi, F. A. (2005). Interaction of Visual and Proprioceptive Feedback During Adaptation of Human Reaching Movements. Journal of Neurophysiology, 93 (6), 3200–3213. 10.1152/jn.00947.2004 Publisher: American Physiological Society

Schroeder, K. E., Irwin, Z. T., Bullard, A. J., Thompson, D. E., Bentley, J. N., Stacey, W. C., Patil, P. G., & Chestek, C. A. (2017). Robust tactile sensory responses in finger area of primate motor cortex relevant to prosthetic control. Journal of Neural Engineering, 14 (4), 046016. 10.1088/1741-2552/aa7329 Publisher: IOP Publishing

Scott, S. H., & Kalaska, J. F. (1997). Reaching Movements With Similar Hand Paths But Different Arm Orientations. I. Activity of Individual Cells in Motor Cortex [Publisher: American Physiological Society]. Journal of Neurophysiology, 77 (2), 826–852. 10.1152/jn.1997.77.2.826

Seki, K., & Fetz, E. E. (2012). Gating of Sensory Input at Spinal and Cortical Levels during Preparation and Execution of Voluntary Movement. Journal of Neuroscience, 32 (3), 890–902. 10.1523/JNEUROSCI.4958-11.2012 Publisher: Society for Neuroscience Section: Articles

Sergio, L. E., & Kalaska, J. F. (2003). Systematic Changes in Motor Cortex Cell Activity With Arm Posture During Directional Isometric Force Generation [Publisher: American Physiological Society]. Journal of Neurophysiology, 89 (1), 212–228. 10.1152/jn.00016.2002

Shelchkova, N. D., Downey, J. E., Greenspon, C. M., Okorokova, E. V., Sobinov, A. R., Verbaarschot, C., He, Q., Sponheim, C., Tortolani, A. F., Moore, D. D., Kaufman, M. T., Lee, R. C., Satzer, D., Gonzalez-Martinez, J., Warnke, P. C., Miller, L. E., Boninger, M. L., Gaunt, R. A., Collinger, J. L., … Bensmaia, S. J. (2023). Microstimulation of human somatosensory cortex evokes task-dependent, spatially patterned responses in motor cortex. Nature Communications, 14, 7270. 10.1038/s41467-023-43140-2

Shenoy, K. V., Sahani, M., & Churchland, M. M. (2013). Cortical Control of Arm Movements: A Dynamical Systems Perspective. Annual Review of Neuroscience, 36 (1), 337–359. 10.1146/annurev-neuro-062111-150509 eprint: https://doi.org/10.1146/annurev-neuro-062111-150509

Sober, S. J., & Sabes, P. N. (2003). Multisensory Integration during Motor Planning. Journal of Neuroscience, 23 (18), 6982–6992. 10.1523/JNEUROSCI.23-18-06982.2003 Publisher: Society for Neuroscience Section: Behavioral/Systems/Cognitive

Suminski, A. J., Tkach, D. C., Fagg, A. H., & Hatsopoulos, N. G. (2010). Incorporating Feedback from Multiple Sensory Modalities Enhances Brain–Machine Interface Control. Journal of Neuroscience, 30 (50), 16777–16787. 10.1523/JNEUROSCI.3967-10.2010

Suminski, A. J., Tkach, D. C., & Hatsopoulos, N. G. (2009). Exploiting multiple sensory modalities in brain-machine interfaces. Neural Networks: The Official Journal of the International Neural Network Society, 22 (9), 1224–1234. 10.1016/j.neunet.2009.05.006

Suresh, A. K., Goodman, J. M., Okorokova, E. V., Kaufman, M., Hatsopoulos, N. G., & Bensmaia, S. J. (2020). Neural population dynamics in motor cortex are different for reach and grasp (S. R. Santacruz, R. B. Ivry, S. R. Santacruz, & M. Capogrosso, Eds.). eLife, 9, e58848. 10.7554/eLife.58848 Publisher: eLife Sciences Publications, Ltd

Velliste, M., Perel, S., Spalding, M. C., Whitford, A. S., & Schwartz, A. B. (2008). Cortical control of a prosthetic arm for self-feeding [Number: 7198 Publisher: Nature Publishing Group]. Nature, 453 (7198), 1098–1101. 10.1038/nature06996

Wang, W., Chan, S. S., Heldman, D. A., & Moran, D. W. (2007). Motor Cortical Representation of Position and Velocity During Reaching [Publisher: American Physiological Society]. Journal of Neurophysiology, 97 (6), 4258–4270. 10.1152/jn.01180.2006

Weber, D. J., London, B. M., Hokanson, J. A., Ayers, C. A., Gaunt, R. A., Torres, R. R., Zaaimi, B., & Miller, L. E. (2011). Limb-state information encoded by peripheral and central somatosensory neurons: Implications for an afferent interface. IEEE transactions on neural systems and rehabilitation engineering: a publication of the IEEE Engineering in Medicine and Biology Society, 19 (5), 501–513. 10.1109/TNSRE.2011.2163145

Wolpert, D. M., & Ghahramani, Z. (2000). Computational principles of movement neuroscience. Nature Neuroscience, 3 (11), 1212. 10.1038/81497

Zhang, J., Riehle, A., Requin, J., & Kornblum, S. (1997). Dynamics of Single Neuron Activity in Monkey Primary Motor Cortex Related to Sensorimotor Transformation. Journal of Neuroscience, 17 (6), 2227–2246. 10.1523/JNEUROSCI.17-06-02227.1997

